# Impact of LP.8.1-Adapted mRNA Vaccination on SARS-CoV-2 Variant Neutralisation

**DOI:** 10.1101/2025.10.21.25338461

**Authors:** C Happle, M Hoffmann, MV Stankov, I Nehlmeier, A Eichmann, T Witte, L Manthey, S Pöhlmann, A Dopfer-Jablonka, GMN Behrens

**Author notes:** Corresponding author: Georg M.N. Behrens, Department of Rheumatology and Immunology, Hannover Medical School, Carl-Neuberg-Straße 1, D - 30625 Hannover, Germany, 5324. co-first authorship (C Happle, M Hoffmann). co-senior authorship.

## Abstract

SARS-CoV-2 continues to evolve, with successive variants evading immunity established through prior infection or vaccination. By mid-2025, the XFG lineage emerged and began replacing LP.8.1 across multiple geographical regions, indicating further adaptive evolution within the JN.1-derived clade. We therefore assessed immune responses in 42 healthcare workers who received 30 μg of LP.8.1-adapted mRNA vaccine (Comirnaty LP.8.1, BioNTech–Pfizer, Mainz, Germany) in September 2025. Post LP.8.1 vaccination, anti-spike IgG and anti-spike omicron IgG changed 2·2-fold and 1·9-fold, respectively, revealing significant increases. Neutralising antibody responses against pseudovirus particles (pp) with spike proteins of various SARS-CoV-2 variants revealed a significant increase in neutralisation of JN.1pp (mean change 4·2-fold), LP.8.1pp (8·2-fold), NB.1.8.1pp (8·6-fold), XFGpp (16·5-fold), and BA.3.2.2pp (2·2-fold). Fold increase in GMT neutralisation post-vaccination was highest for XFGpp whilst absolute post-vaccination neutralisation GMT was lowest for BA.3.2.2pp. Our data suggest that the LP.8.1 mRNA vaccination most likely increases protection against severe disease courses and sequelae of COVID-19 caused by the currently circulating XFG variant. Strengthening humoral immunity against BA.3.2 variants may require further vaccine refinement.

---

SARS-CoV-2 continues to evolve, with successive variants evading immunity established through prior infection or vaccination. In mid-2024, a vaccine tailored to the JN.1 variant was authorized, which boosted neutralizing antibody responses and provided substantial protection against severe disease and hospitalization (1-3). Around six months later, in January 2025, LP.8.1, a JN.1 descendant, was classified as a *variant under monitoring* by the World Health Organization, due to its epidemiological significance and enhanced transmission fitness relative to contemporaneous strains. After emerging in late 2024, LP.8.1 rapidly overtook the XEC variant, establishing dominance throughout the Americas and Europe by early 2025. In the United States, LP.8.1 represented approximately 50% of COVID-19 cases by May 2025 (4). While KP.2- and JN.1-adapted vaccines generated neutralising antibodies against LP.8.1, these titres were reduced compared to earlier JN.1-lineage variants, indicating continued antigenic drift (5). By mid-2025, the XFG lineage emerged and began replacing LP.8.1 across multiple geographical regions, indicating further adaptive evolution within the JN.1-derived clade. Subsequent investigations confirmed robust immune evasion coupled with diminished ACE2 receptor binding efficiency for the XFG variant (5).

Given antigenic evolution and declining neutralising antibody concentrations, protection conferred by the JN.1-adapted vaccine is now considered reduced. Consequently, on July 24, 2025, the European Medicines Agency (EMA) authorized a monovalent mRNA COVID-19 vaccine encoding the LP.8.1 variant spike protein (Comirnaty LP.8.1, BioNTech–Pfizer, Mainz, Germany). This formulation represents the most current update to the mRNA vaccine platform, designed to target the prevalent circulating strain.

Upon authorization, data on human immunogenicity and clinical effectiveness remained scarce. We therefore assessed immune responses in 42 healthcare workers (median age 57 years [IQR 16]; 18 [43%] male) who received 30 μg of the LP.8.1-adapted mRNA vaccine in September 2025. We obtained serum samples immediately before vaccination and at a median of 14 days (IQR 0, range 13– 16 days) post-vaccination to characterise humoral immune responses. Participants had received a median of five previous COVID-19 vaccinations (IQR 1, range 3–9), with a median interval of 13 months (IQR 8, range 10–46 months) since their last vaccination. Of 42 individuals, 35 (83%) reported at least one prior SARS-CoV-2 infection (median 1 infection [IQR 1, range 0–5], Appendix p2).

We first quantified SARS-CoV-2 anti-spike IgG antibodies before and following LP.8.1 vaccination. Prior to LP.8.1 immunisation, participants showed a median of 1994·0 (IQR 1910·0) binding-antibody units (BAU) per mL of anti-spike (Wuhan-Hu-S1) IgG antibodies and a median of 422·1 (IQR 238·2) relative units (RU) per mL of anti-spike omicron IgG antibodies **(Figure A)**. Post LP.8.1 vaccination, anti-spike IgG and anti-spike omicron IgG changed 2·2-fold and 1·9-fold, respectively, revealing significant increases (p<0·0001; Figure A) comparable to that observed after vaccination with the JN.1-adapted vaccine (3).

Next, we assessed neutralising antibody responses against pseudovirus particles (pp) with spike proteins of various SARS-CoV-2 variants before and after vaccination. Pre-vaccination sera demonstrated neutralising activity against JN.1pp (response rate 100%, geometric mean titre [GMT] 467), LP.8.1pp (95%, GMT 169), NB.1.8.1pp (100%, GMT 126), XFGpp (81%, GMT 24), and BA.3.2.2pp (100%, GMT 92). Compared with LP.8.1pp, pre-vaccination neutralisation was 2·8-fold higher for JN.1pp, but lower for the others with a 1·3-fold change for NB.1.8.1pp, 7·0-fold change for XFGpp, and a 1·8-fold change in neutralization for BA.3.2.2pp **(Figure B)**. After vaccination, the response rates increased significantly for all pseudoviruses **(Figure C)**. We observed a significant increase in neutralisation of JN.1pp (mean change 4·2-fold), LP.8.1pp (8·2-fold), NB.1.8.1pp (8·6-fold), XFGpp (16·5-fold), and BA.3.2.2pp (2·2-fold). Fold increase in GMT neutralisation post-vaccination was highest for XFGpp whilst absolute post-vaccination neutralisation GMT was lowest for BA.3.2.2pp.

XFG is currently the most prevalent variant on the global level and the LP.8.1 vaccination strongly augments the neutralisation for this variant. However, we also found that BA.3.2, which has spread across four continents with low prevalence, evades antibody responses after vaccination with higher efficiency than the globally dominant variant XFG (6,7). Consequently, the neutralisation potential installed upon LP.8.1 vaccination may not be sufficient to halt BA.3.2 from achieving high global prevalence in the future, although additional factors including improved ACE2 binding and augmented lung cell entry are likely prerequisites for such expansion (7).

We note that our study population exhibited rates of hybrid immunity against SARS-CoV-2 variants before vaccination, with 83% experiencing prior SARS-CoV-2 infections and all being previously vaccinated against SARS-CoV-2 with a median of five vaccinations. This could have affected the magnitude and quality of humoral immunity induced by the LP.8.1-adapted vaccine and might not be representative of other populations. Despite these considerations, and some limitations listed in detail in the Appendix (p 5), our data support the notion that the mRNA vaccine against omicron LP.8.1 most likely increases protection against severe disease courses and sequelae of COVID-19 caused by the currently circulating XFG variant. Strengthening humoral immunity against BA.3.2 variants may require further vaccine refinement.

## Data Availability

All data produced in the present study are available upon reasonable request to the authors

---

**Figure.**
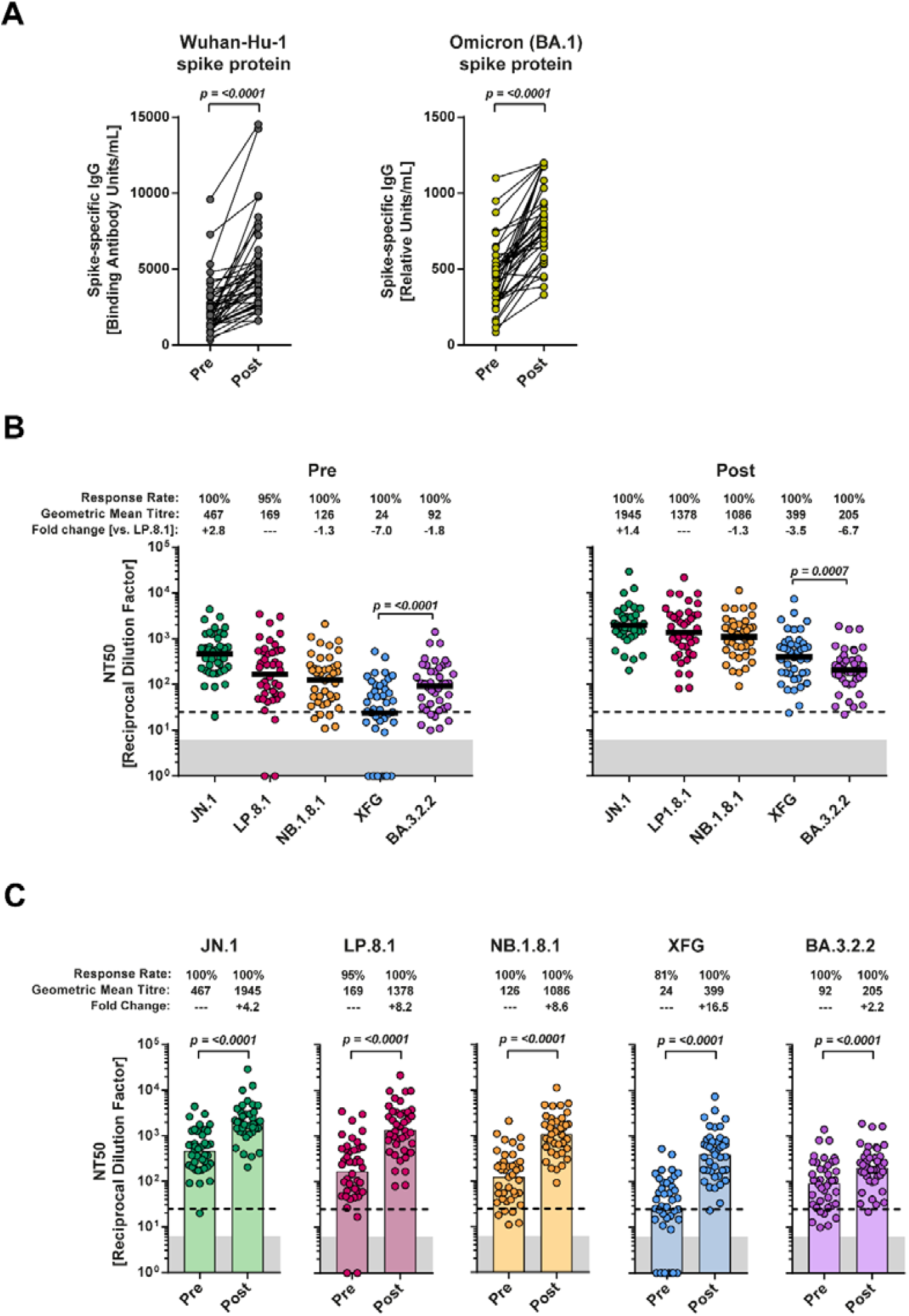
Humoral immune responses after mRNA omicron LP.8.1 vaccination. **(A)** Serum concentrations of Wuhan-Hu-1 spike-specific IgG and omicron S-specific IgG (n=42) obtained before (Pre) or after (Post) vaccination with the mRNA omicron LP8.1 vaccine. **(B)** Neutralisation of vesicular stomatitis virus-based pseudovirus particles bearing the indicated S proteins by donor-matched serum (n=41). Data are grouped to compare differences in SARS-CoV-2 lineage-specific neutralisation before and after omicron LP.8.1 vaccination. Information on GMT (also indicated by horizontal lines), response rates, and mean fold change in neutralisation compared with LP.8.1 pseudovirus particles are indicated above the graphs. Data represent GMT (coloured columns) from a single experiment, performed with four technical replicates. The lowest serum dilution tested (dashed lines) and the threshold (lower limit of detection; grey shaded areas) are indicated. Of note, for graphical reasons, serum samples yielding a 50% neutralisation titre value below 6·25 (limit of detection) were manually set at bottom of the axis. **(C)** Data are regrouped to compare differences in SARS-CoV-2 lineage-specific neutralization before and after vaccination. Information on response rates and mean fold change in neutralisation after vaccination are indicated above the graphs. Individual neutralisation data are available in the appendix (pp 9–10). GMT=geometric mean titres.

