## Appendix for "Impact of LP.8.1-Adapted mRNA Vaccination on SARS-CoV-2 Variant Neutralisation"

### Content

Table………………………………………………………………….. 2

Methods…………………………………………………………………. 3

Limitations of the study……………………………………………….. 5

Acknowledgement……………………………………………………….. 6

Supplementary references……………………………………………... 7

Supplementary figures………………………………………………… 8

**Table S1. Demographics and infection and vaccination history**

| Variable | Vaccinees |
|---|---|
| Number of vaccinees | 42 |
| Age, Median (IQR) | 57 (16) |
| Sex, male (%) | 18 (43) |
| Prior COVID-19 vaccination (%) | 42 (100) |
| Number of COVID-19 vaccinations[1]: Median (IQR), range | 5 (1), 3-9 |
| Months since last COVID-19 vaccination[2]: Median (IQR), range | 13 (8), 10-46 |
| Prior SARS-CoV-2 infection, n (%) | 35 (83) |
| Number of SARS-CoV-2 infections: Median (IQR), range | 1 (1), 0-5 |
| Months since last SARS-CoV-2 infection[3]: Median (IQR), range | 27 (18), 8-48 |

[1] Missing information for one person. [2] Missing information for three persons. [3] Missing information for five persons

### Methods

*Participants*

We analysed n=42 individuals vaccinated with Corminaty LP.8.1 from the COVID-19 Contact (CoCo) Study (German Clinical Trial Registry, DRKS00021152), an ongoing, prospective, observational study monitoring anti-SARS-CoV-2 immunoglobulin G and immune responses in healthcare professionals at Hannover Medical School [1].

Our sample size calculation indicated that n=42 participants would provide adequate statistical power to identify a clinically significant within-group difference. This calculation assumed that anti-S protein IgG concentrations would increase twofold following vaccination (baseline: mean 822 RU/mL (SD 747); post-vaccination: mean 1,644 RU/mL (SD 1,494)), with an inter-group correlation of 0.5. We based these assumptions on anti-S IgG data obtained from a convenience sample of 24 individuals within the CoCo cohort during August 2023. The calculation utilised a one-tailed paired t-test comparing mean differences, with 95% statistical power and 1% significance level. Accounting for an anticipated ca. 20% attrition rate during the study timeframe, we determined that enrolling 52 vaccinated individuals would be adequate. We conducted the power analysis using G*Power software, Version 3.1.9.6.

Participants that were vaccinated (n=52) in September 2025 with Corminaty LP.8.1 (30µg) were invited to donate blood before and two weeks after vaccination at which time robust antibody responses are detectable[2] . The CoCo Study cohort comprises a general population of mainly health-care professionals. For n=43 participants, blood samples before and after vaccination (day 14) were available. One individual developed positive anti-NCP IgG after vaccination and was excluded from the analysis, all other reported no COVID-19 after vaccination and did not turn positive for anti-NCP IgG. The remaining n=42 participants went into the final analysis for the ELISA measurements, and we assessed n=41 participants for neutralisation, since one post-vaccination sample was unavailable for testing. Forty-eight percent of the LP.8.1 vaccinees reported underlying conditions, with cardiovascular (29%) and respiratory diseases (7%) most prevalent morbidities. Five people (12%) reported intake of immunosuppressive drugs without further specification. Follow-up sampling was performed two weeks (median 14 days, IQR 0, range 13-16 days) after LP.8.1 vaccination. Demographics (sex and age), and infection, and vaccination history, respectively, are depicted in **Table S1**.

*Serology*

We separated serum after venous blood collection and stored it at -20 °C until use. We measured SARS-CoV-2 Spike IgG by quantitative ELISA (anti-SARS-CoV-2 S1 Spike protein [Wuhan-Hu-S1] domain/receptor binding domain IgG [SARS-CoV-2-QuantiVac, EI 2606-9601-10 G], and anti-SARS-CoV-2 S1 Spike protein (Omicron domain/receptor binding domain IgG [EI 2606-9601-30 G, both EUROIMMUN, Lübeck, Germany) according to the manufacturer's instructions (dilution up to 1:4,000). We express anti-Omicron Spike IgG concentrations as RU/mL and anti-Wuhan-Hu-S1 Spike results as binding antibody units (BAU/mL). We performed anti-SARS-CoV-2 nucleocapsid (NCP) IgG measurements according to the manufacturer's instructions (EI 2606-9601-2 G, EUROIMMUN, Lübeck, Germany). All ELISAs were measure on a VARIOSKAN LUX microplate reader (Thermo Scientific, Vantaa, Finland). Instrument control and primary data acquisition were performed utilizing SkanIt RE software, version 6.1.1 (Thermo Scientific). Subsequent statistical analysis, including curve fitting, comparison of means, and graphical representation, was conducted using GraphPad Prism, version 9 (GraphPad Software, San Diego, California).

*Production of pseudovirus particles and pseudovirus neutralisation test (pVNT)*

Production of pseudovirus particles bearing SARS-CoV-2 spike proteins was carried based on a previously protocol established for the MERS-CoV spike protein[3]. First, 293T cells were transfected by calcium phosphate precipitation with expression plasmids for the respective spike proteins or empty expression vector (negative control). The next day, the cell culture supernatant was aspirated and cells were washed once with PBS, before being inoculated with VSV-G-trans complemented VSV*ΔG (FLuc)[4] at a multiplicity of infection of 3. The original VSV*ΔG (FLuc) stock was kindly provided by Gert Zimmer). Following an incubation time of 1h, the supernatant was aspirated and cells were washed twice with PBS, before medium containing anti-VSV-G antibody (1:1000-diluted supernatant from I1-hybridoma cells; ATCC CRL-2700) was added and cells were further incubated for 18-20 h. After this incubation period, the pseudovirus particle-containing supernatants were harvested and centrifuged (4,000 x g for 10 min at room temperature) to remove cellular debris. Finally, clarified supernatants were aliquoted and stored at -80 °C until further use. Of note, quality of the pseudovirus particles was assessed before their use in neutralisation experiments by inoculation of Vero cells and measurement of infection efficiency by luciferase assay (procedure is stated below below). Expression plasmids for spike proteins of the following SARS-CoV-2 lineages were used: JN.1 (hCoV-19/Denmark/DCGC-663536/2023, EPI_ISL_18530042), LP.8.1 (hCoV-19/Canada/ON-KHS-05478-v1/2024, EPI_ISL_19553000), NB.1.8.1 (hCoV-19/Singapore/Y25R15MSC85/2025, EPI_ISL_19826033), XFG (hCoV-19/Spain/PV-HUD-39393424/2025, EPI_ISL_20053327), and BA.3.2.2 (hCoV-19/Germany/NW-RKI-I-1148174/2025, EPI_ISL_19893762). All spike protein sequences were codon-optimised, contained a C-terminal truncation of 18 amino acid residues and were cloned into the pCG1 expression plasmid that was kindly provided by Roberto Cattaneo (Department of Molecular Medicine, Mayo Clinic, Rochester, Minnesota, USA). Further, spike protein sequences were verified by Sanger sequencing using a commercial service (Microsynth SeqLab). Information on SARS-CoV-2 lineages and corresponding mutations was retrieved from the GISAID (https://gisaid.org/) and CoV-Spectrum (https://cov-spectrum.org/) databases.

The pseudovirus particle neutralization assay (pVNT) was carried out based on a previously described protocol[5]. First, pseudovirus particles were mixed with equal volumes of serially diluted serum (serum was diluted in culture medium) or culture medium without serum (no serum control). In total, six serum dilutions (final dilution after mixing with pseudovirus particles: 1:25, 1:100, 1:400, 1:1600, 1:6,400, 1:12,800) and the respective no serum control were tested per sample and all samples were tested with four technical replicates. After mixing, samples were incubated at for 30 min before they were added to Vero cells grown in 96-well plates. Following an incubation time of 16-18h, the cell culture supernatant was aspirated and cells were lysed by incubation (30 min at room temperature) with PBS containing 0.5% Tergitol (50 µL/well). Next, cell lysates were transferred into white 96-well plates and mixed with and equal volume of firefly luciferase substrate (Beetle-Juice, PJK), before luminescence was recorded using a Hidex Sense microplate reader (Hidex). Neutralisation efficiency was further determined based on the relative reduction of luciferase activity in serum-containing samples compared samples that did not contain serum (set as 0% neutralisation). Finally, dose-response curves were generated based on a non-linear regression model and 50% neutralising titres (NT50) were determined for each serum-pseudovirus combination. Of note, serum samples that did not yield an NT50 value of at least 6.25 (25% of the lowest serum dilution analysed) were considered negative and assigned an NT50 value of 1.

*Statistics*

Statistical analysis was conducted using GraphPad Prism 8.4 or 9.0 (GraphPad Software, USA). Outliers were included in the analysis, and missing values were excluded pairwise. Median (IQR) was used for non-normally distributed data and Wilcoxon-Mann-Whitney-tests were used for comparison. Neutralisation titres were transformed to geometric mean titres.

**Limitations of the study**

Our study has some limitations. Although our previous analysis[2] confirmed that anti-S IgG and neutralising antibodies plateaued at day eight to ten post Omicron XBB.1.5 vaccination, which is in keeping with antibody kinetics described after the second BNT162b2 vaccination[6] or after other COVID-19 vaccinations[7], our data are preliminary and humoral immunity could further strengthen over time. Whilst our study was powered to demonstrate clinically meaningful changes in immune surrogates after COVID-19 vaccination (anti-spike IgG), overall sample size is small and limits interpretation. Thus, this data can only provide first insights into the initial immune response to the updated LP.8.1 vaccine. Longitudinal data will be necessary to assess immune trends and durability. In addition, most of our vaccinees had previous SARS-CoV-2 Omicron infections and vaccinations, contributing to considerable S protein responsive already before LP.8.1 vaccination. In regards to comparisons with previous reports, we would like to point out that the vaccinees described here were partially different (e.g. mean age) compared to participants described in our previous studies about the XBB.1.5[2] and JN.1 vaccination (Ref. 3 and 7, main manuscript). Only 50% of the current participants took part in studies described in Ref. 3 and 7 and 28% did not receive a JN.1 vaccination. Finally, neutralisation of SARS-CoV-2 lineages was assessed by pVNT, which has been shown to serve as an adequate surrogate model for this purpose[8]. Nevertheless, our data formally await confirmation with clinical isolates and eventually validation in studies with clinical endpoints.

**Acknowledgements**

**Contributions**
Study design: G.M.N.B., A.D.-J.
Data collection: I.N., A.E., L.M., C.H., M.V.S., T.W.
Data curation: C.H., M.V.S
Data analysis: M.H, C.H., L.M, M.V.S, G.M.N.B., T.W.
Data interpretation: M.H., S.P., A.D.-J., G.M.N.B.
Writing: G.M.N.B. with comments from all authors.
G.M.N.B, M.H., and A.D.-J. have directly accessed and verified the underlying data reported in the manuscript.

**Data sharing statement**
All requests for raw and analysed data that underly the results reported in this article will be reviewed within four weeks by the CoCo Study Team, Hannover Medical School to determine whether the request is subject to confidentiality and data protection obligations. Data that can be shared will be released via a material transfer agreement.

**Ethics committee approval**
The CoCo Study (German Clinical Trial Register DRKS00021152) and the analysis conducted for this article were approved by the Internal Review Board of Hannover Medical School (institutional review board no. 8973_BO-K_2020, last amendment Aug 2024). All study participants gave written informed consent and received no compensation.

**Competing Interests and Funding**
S.P., M.H., I.N., and A.E. declare conducting contract research (testing of vaccinee sera for neutralising activity against SARS-CoV-2) for Valneva, unrelated to this work, and S.P. served as an advisor for BioNTech, unrelated to this work. G.M.N.B. declares serving as a lecturer for Pfizer and adviser for Moderna, unrelated to this work. A.D.-J. declares serving as an advisor for Pfizer, unrelated to this work. G.M.N.B. and A.D.-J. acknowledge funding (COVID-19-Research Network Lower Saxony (COFONI) through funding from the Ministry of Science and Culture of Lower Saxony in Germany (14-76103-184, project 4LZF23), G.M.N.B. acknowledge funding by the European Regional Development Fund ZW7-85151373, and A.D.-J. acknowledge funding by European Social Fund (ZAM5-87006761). T.W. acknowledges funding by the German Research Foundation (Deutsche Forschungsgemeinschaft, DFG) under Germany's Excellence Strategy EXC 2155 RESIST (project 390874280). S.P. acknowledges funding by the EU project UNDINE (grant agreement number 101057100), the COVID-19-Research Network Lower Saxony (COFONI) through funding from the Ministry of Science and Culture of Lower Saxony in Germany (14-76403-184, projects 7FF22, 6FF22, 10FF22), The Federal Ministry of Education and Research (COVIM 2.0, 01KX2121), and the German Research Foundation (Deutsche Forschungsgemeinschaft, DFG; PO 716/11-1). The funding sources had no role in the design and execution of the study, the writing of the manuscript and the decision to submit the manuscript for publication. The authors did not receive payment by a pharmaceutical company or other agency to write the publication. The authors were not precluded from accessing data in the study, and they accept responsibility to submit for publication.

We thank the CoCo Study participants for their support and the entire CoCo study team for help. We would like to thank J. Topal, K. Sträche, B. Heinisch, R. Rubin, S. Ritter, H. May, and A. Stölting for technical and logistical support.

**Suppl. References**

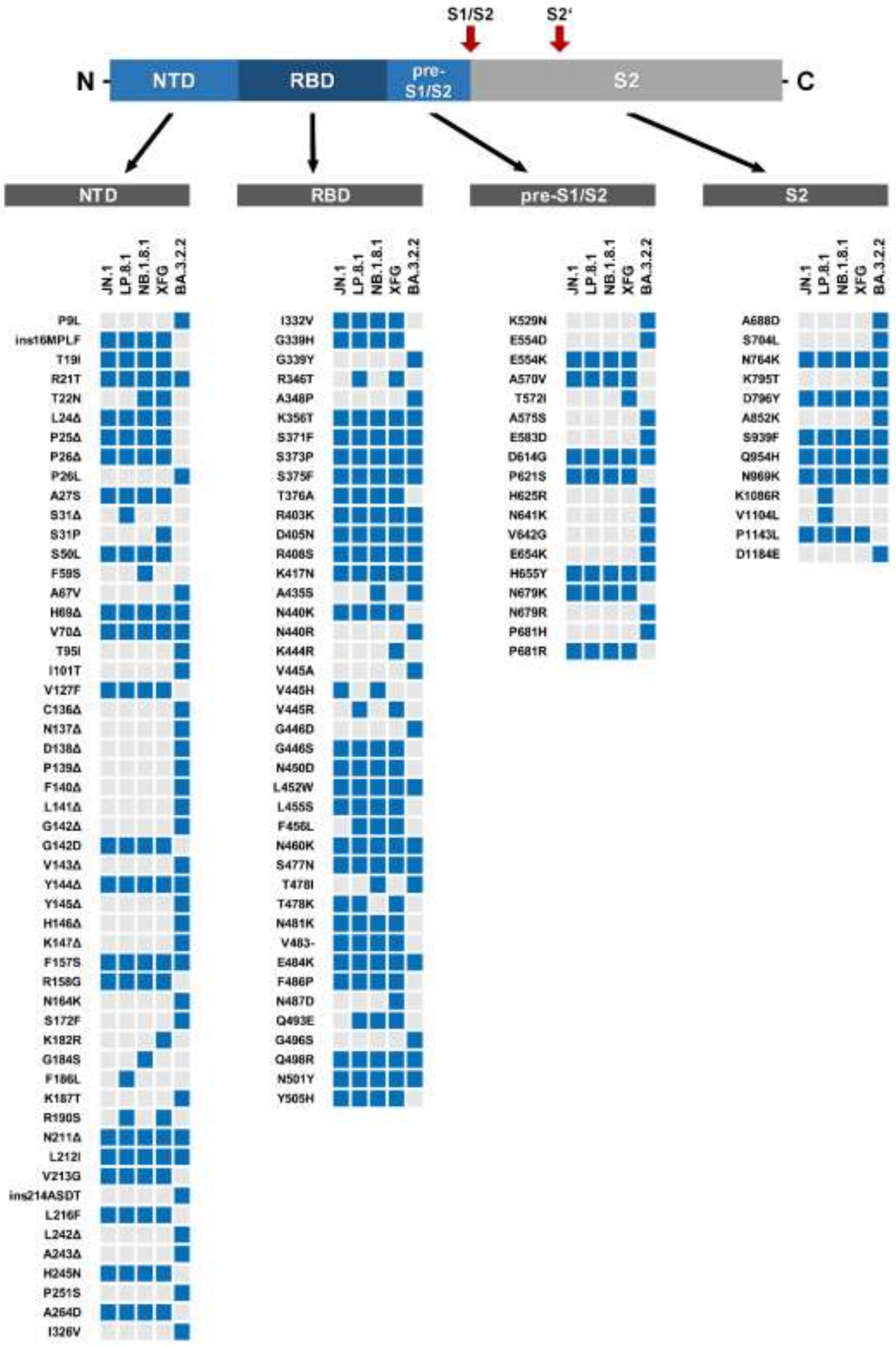


**Fig. S1 | Overview on SARS-CoV-2 lineage-specific spike protein mutations.**

Top: Schematic representation of the SARS-CoV-2 spike protein with domains and cleavage site (S1/S2 and S2') highlighted. Abbreviations: NTD = N-terminal domain, RBD = receptor-binding domain, pre-S1/S2 = region between the RBD and the S1/S2 cleavage site, S2 = S2 subunit. Bottom: Spike protein mutations in JN.1, LP.8.1, NB.1.8.1, XFG, and BA.3.2.2 relative to the Wuhan-Hu-1 isolate are indicated by blue boxes.

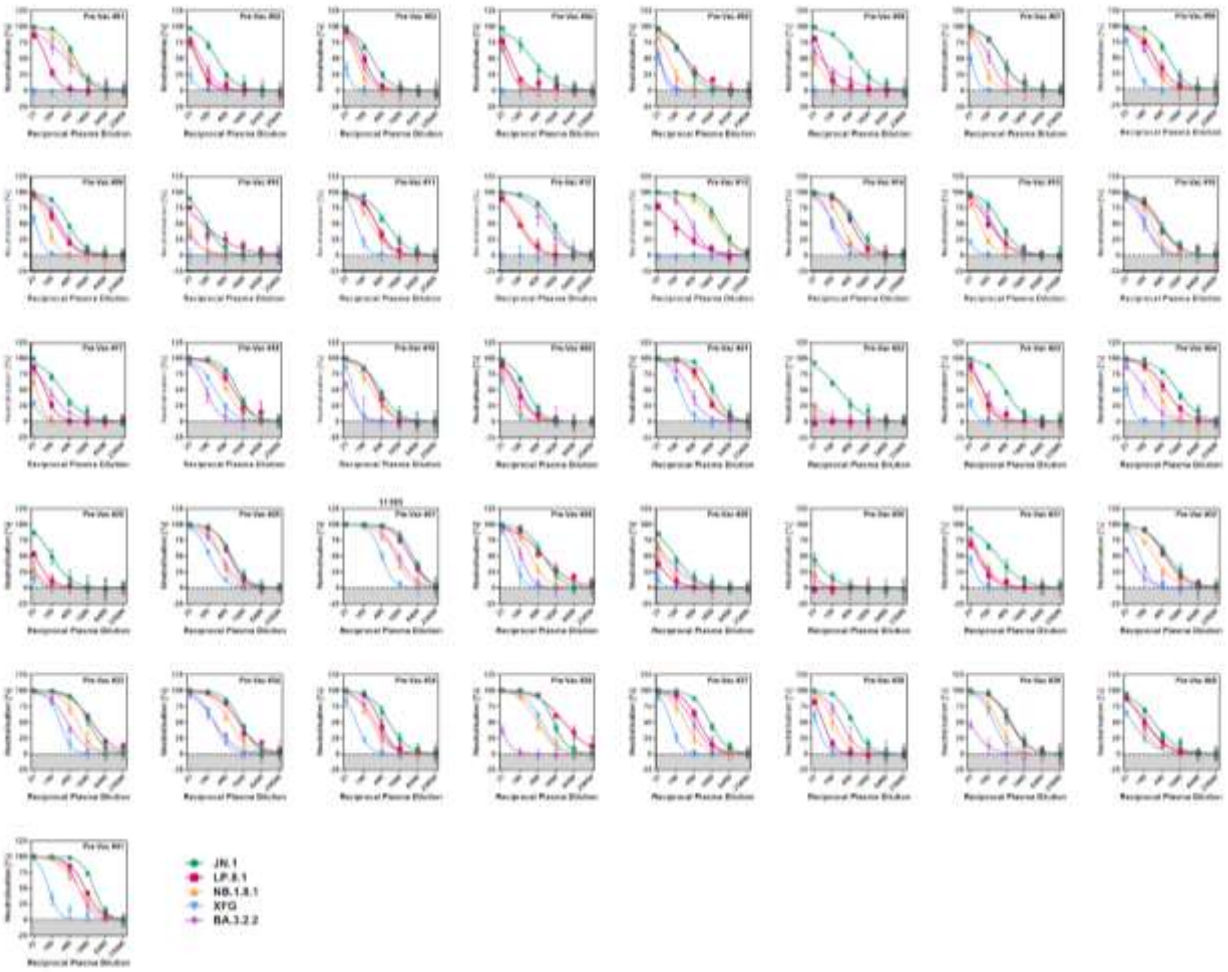


**Fig. S2 | Individual neutralisation data for pre-vaccination serum.**

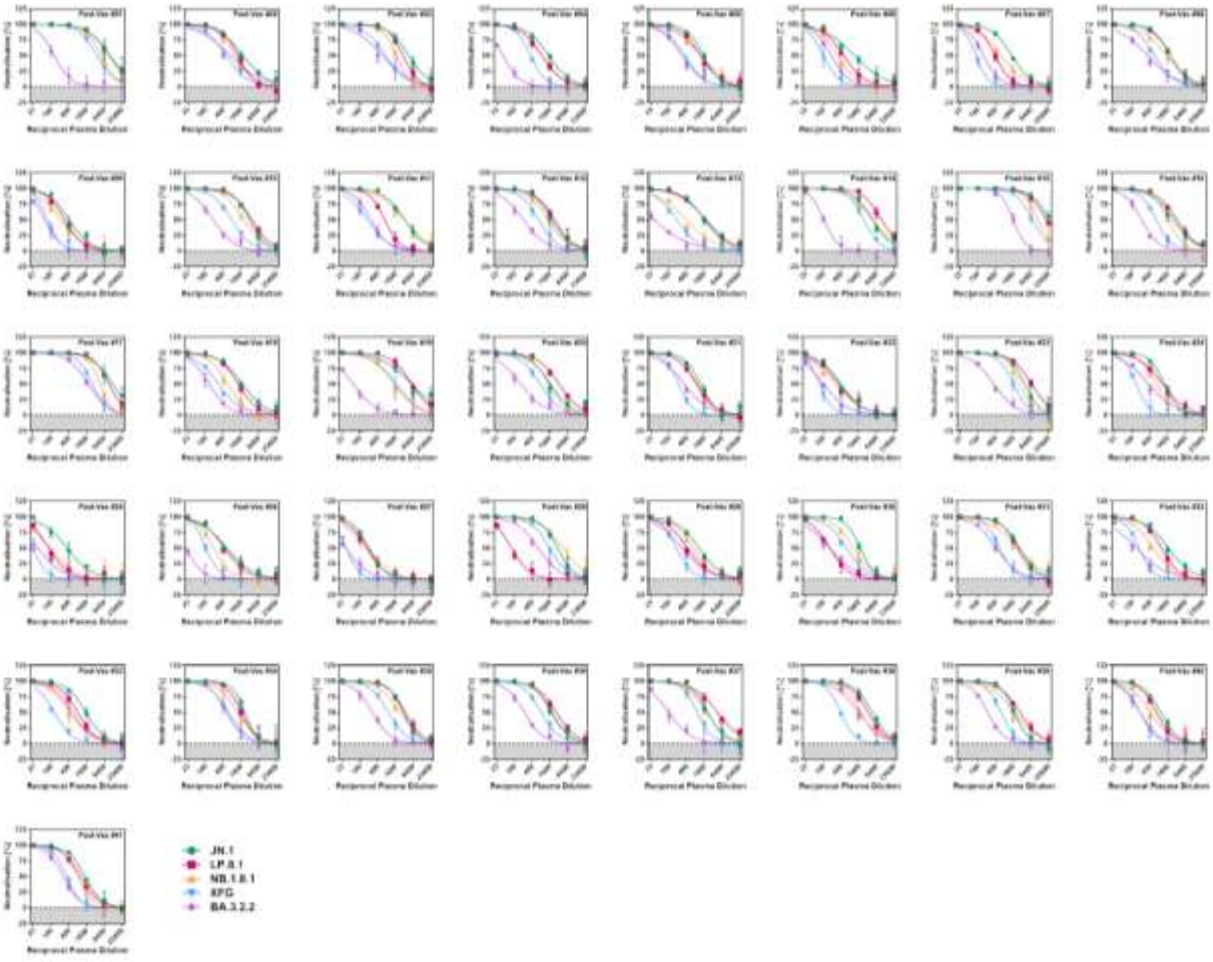


**Fig. S3 | Individual neutralisation data for post-vaccination serum.**
